# Associations of amyloid biomarkers with brain and cognitive changes from imaging, spinal fluid, and plasma

**DOI:** 10.64898/2026.02.05.26345647

**Authors:** Jason Scully, Mahsa Dadar, Cassandra Morrison, the Alzheimer’s Disease Neuroimaging Initiative

**Affiliations:** Department of Psychology, Carleton University, Ottawa, Ontario, K1S 5B6, Canada; Department of Psychiatry, McGill University, Montreal, Quebec, H3A 1A1, Canada; Douglas Mental Health University Institute, Montreal, Quebec, H4H 1R3, Canada

**Keywords:** Amyloid, PET, CSF, Plasma, White Matter Hyperintensities, Cognition, Brain Structures

## Abstract

**BACKGROUND:** Positron emission tomography (PET), cerebrospinal fluid (CSF), and plasma assessments are used to measure amyloid abnormality in Alzheimer’s disease (AD). However, it remains unclear if these three measures are similarly associated with brain structure and cognitive measures.

**METHODS:** Linear regressions examined the relationship between amyloid levels measured by PET, CSF, and plasma and brain volumes, white matter hyperintensities (WMHs), and cognitive measures.

**RESULTS:** Moderate correlations were found between PET and CSF amyloid measurements and PET and plasma measurements, while weak correlations were found between CSF and plasma. PET, CSF, and plasma amyloid measurements differed in their associations with brain volume, WMHs, and cognition.

**DISCUSSION:** Using different measurement methods, amyloid was not consistently associated with volumetric or cognitive measures. Our findings also suggest that plasma markers may not be associated with cognitive and brain changes in the same manner as CSF and PET.

## Introduction

Over the last 50 years, global life expectancy has increased from 58 to 71,^1^ and by 2030, the population aged 60 and over will increase from 1 to 1.4 billion.^2^ With increased age, a surge in health conditions such as hearing loss, diabetes, and dementia occurs.^2^ Dementia has quickly become a global health problem, causing substantial personal, societal, and economic burdens. The most common cause of dementia in aging populations is Alzheimer’s disease (AD),^3^ a neurodegenerative disease characterized by the buildup of beta-amyloid, neurofibrillary tau proteins, and increased brain atrophy (i.e., neurodegeneration).^4^ Amyloid and tau accumulation alongside neurodegeneration leads to cognitive decline and functional impairments.^5^ To measure these AD pathologies (amyloid, tau, and neurodegeneration), brain imaging techniques such as positron emission tomography (PET) and magnetic resonance imaging (MRI) as well as cerebrospinal fluid (CSF) and blood plasma assays are often employed.^4^ Given that neurodegeneration and tau accumulation are also present in other neurodegenerative disorders,^6^ amyloid plaques may be the defining pathological marker of AD to help distinguish it from other dementias.^7^

Currently, there are no safe and effective treatments to cure AD,^8,9^ but new treatments such as lecanemab^10^ and donanemab target amyloid to reduce the buildup of plaques.^11^ Before initiating these treatments, biomarker confirmation of amyloid pathology is typically required. Although the FDA does not specify a diagnostic tool, PET and CSF analyses are the most commonly employed approaches.^11–13^ However, the feasibility of PET and CSF is limited due to the high cost and limited accessibility of PET and invasiveness of CSF collection.^14,15^ For these reasons, research is exploring the practicality of using blood plasma biomarkers as an inexpensive and less invasive alternative to detect amyloid abnormality than PET and CSF.^16^ To our knowledge, the degree to which amyloid PET, CSF, and plasma measurements align with one another has not yet been systematically examined in large samples (see Table 1 for a review of studies). Measurement inconsistency between these modalities (PET, CSF, plasma) may lead to discrepant diagnostic classification and selection of inappropriate treatment options.

**Table 1.**
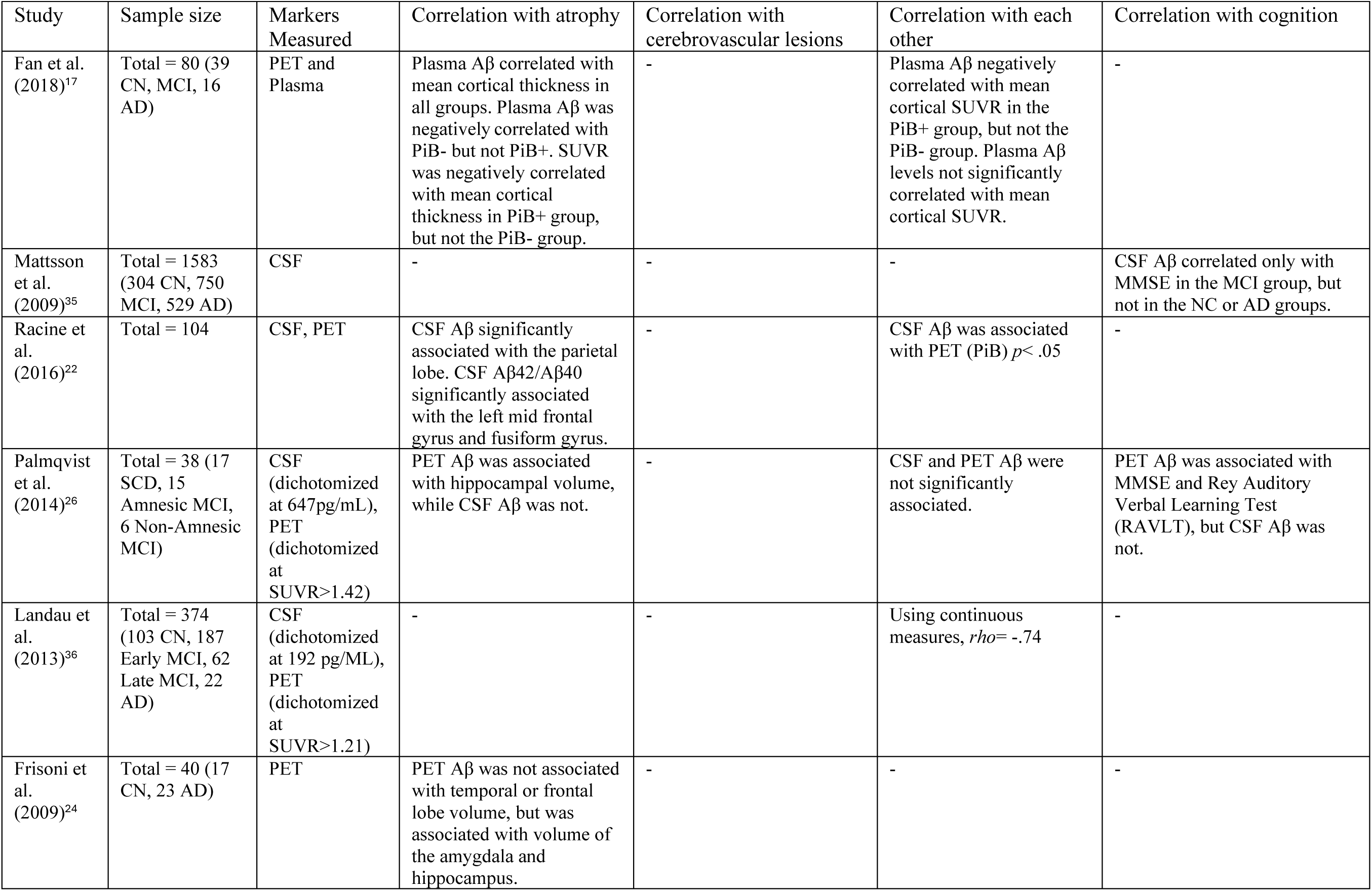
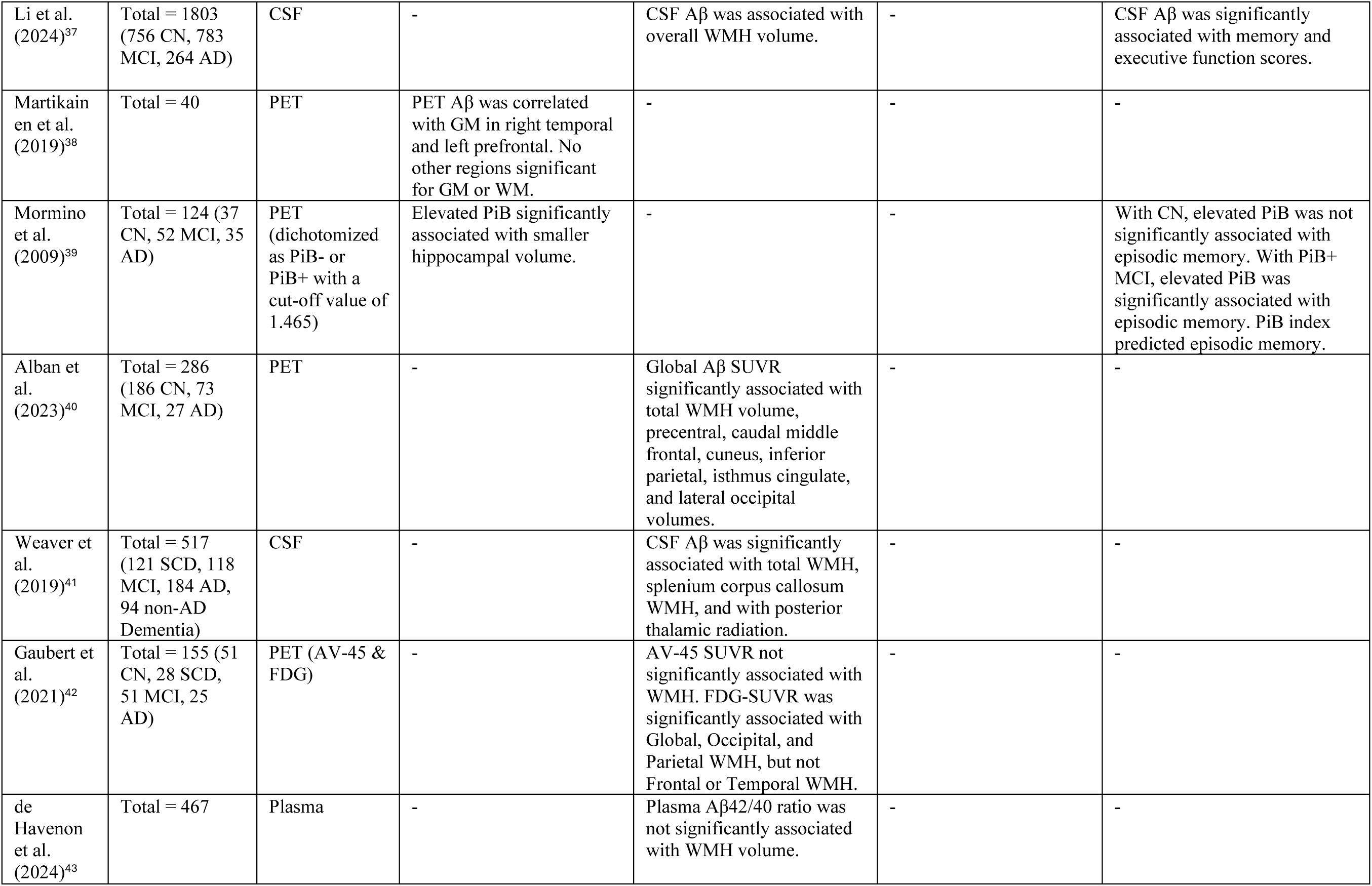
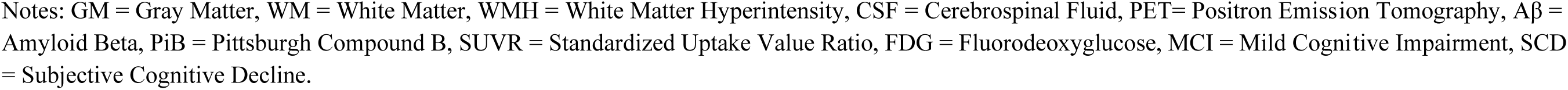
Review of studies investigating the associations of amyloid measurements with atrophy, cerebrovascular lesions, with each other, and with cognition.

| Study | Sample size | Markers Measured | Correlation with atrophy | Correlation with cerebrovascular lesions | Correlation with each other | Correlation with cognition |
| --- | --- | --- | --- | --- | --- | --- |
| Fan et al. (2018)<sup>17</sup> | Total = 80 (39 CN, MCI, 16 AD) | PET and Plasma | Plasma Aβ correlated with mean cortical thickness in all groups. Plasma Aβ was negatively correlated with PiB- but not PiB+. SUVR was negatively correlated with mean cortical thickness in PiB+ group, but not the PiB- group. | - | Plasma Aβ negatively correlated with mean cortical SUVR in the PiB+ group, but not the PiB- group. Plasma Aβ levels not significantly correlated with mean cortical SUVR. | - |
| Mattsson et al. (2009)<sup>35</sup> | Total = 1583 (304 CN, 750 MCI, 529 AD) | CSF | - | - | - | CSF Aβ correlated only with MMSE in the MCI group, but not in the NC or AD groups. |
| Racine et al. (2016)<sup>22</sup> | Total = 104 | CSF, PET | CSF Aβ significantly associated with the parietal lobe. CSF Aβ42/Aβ40 significantly associated with the left mid frontal gyrus and fusiform gyrus. | - | CSF Aβ was associated with PET (PiB) $p < .05$ | - |
| Palmqvist et al. (2014)<sup>26</sup> | Total = 38 (17 SCD, 15 Amnesic MCI, 6 Non-Amnesic MCI) | CSF (dichotomized at 647pg/mL), PET (dichotomized at SUVR>1.42) | PET Aβ was associated with hippocampal volume, while CSF Aβ was not. | - | CSF and PET Aβ were not significantly associated. | PET Aβ was associated with MMSE and Rey Auditory Verbal Learning Test (RAVLT), but CSF Aβ was not. |
| Landau et al. (2013)<sup>36</sup> | Total = 374 (103 CN, 187 Early MCI, 62 Late MCI, 22 AD) | CSF (dichotomized at 192 pg/mL), PET (dichotomized at SUVR>1.21) | - | - | Using continuous measures, $\rho = -.74$ | - |
| Frisoni et al. (2009)<sup>24</sup> | Total = 40 (17 CN, 23 AD) | PET | PET Aβ was not associated with temporal or frontal lobe volume, but was associated with volume of the amygdala and hippocampus. | - | - | - |
| Li et al.<br>(2024)<sup>37</sup> | Total = 1803<br>(756 CN, 783 MCI, 264 AD) | CSF | - | CSF Aβ was associated with overall WMH volume. | - | CSF Aβ was significantly associated with memory and executive function scores. |
| Martikainen et al.<br>(2019)<sup>38</sup> | Total = 40 | PET | PET Aβ was correlated with GM in right temporal and left prefrontal. No other regions significant for GM or WM. | - | - | - |
| Mormino et al.<br>(2009)<sup>39</sup> | Total = 124 (37 CN, 52 MCI, 35 AD) | PET (dichotomized as PiB- or PiB+ with a cut-off value of 1.465) | Elevated PiB significantly associated with smaller hippocampal volume. | - | - | With CN, elevated PiB was not significantly associated with episodic memory. With PiB+ MCI, elevated PiB was significantly associated with episodic memory. PiB index predicted episodic memory. |
| Alban et al.<br>(2023)<sup>40</sup> | Total = 286<br>(186 CN, 73 MCI, 27 AD) | PET | - | Global Aβ SUVR significantly associated with total WMH volume, precentral, caudal middle frontal, cuneus, inferior parietal, isthmus cingulate, and lateral occipital volumes. | - | - |
| Weaver et al.<br>(2019)<sup>41</sup> | Total = 517<br>(121 SCD, 118 MCI, 184 AD, 94 non-AD Dementia) | CSF | - | CSF Aβ was significantly associated with total WMH, splenium corpus callosum WMH, and with posterior thalamic radiation. | - | - |
| Gaubert et al.<br>(2021)<sup>42</sup> | Total = 155 (51 CN, 28 SCD, 51 MCI, 25 AD) | PET (AV-45 & FDG) | - | AV-45 SUVR not significantly associated with WMH. FDG-SUVR was significantly associated with Global, Occipital, and Parietal WMH, but not Frontal or Temporal WMH. | - | - |
| de Havenon et al.<br>(2024)<sup>43</sup> | Total = 467 | Plasma | - | Plasma Aβ42/40 ratio was not significantly associated with WMH volume. | - | - |

Notes: GM = Gray Matter, WM = White Matter, WMH = White Matter Hyperintensity, CSF = Cerebrospinal Fluid, PET= Positron Emission Tomography, $A\beta$ = Amyloid Beta, PiB = Pittsburgh Compound B, SUVR = Standardized Uptake Value Ratio, FDG = Fluorodeoxyglucose, MCI = Mild Cognitive Impairment, SCD = Subjective Cognitive Decline.

Amyloid build-up is positively associated with neurodegeneration,^17^ cerebrovascular pathology (e.g., white matter hyperintensities; WMHs),^18,19^ and cognitive decline.^20,21^ Studies using CSF amyloid have shown associations with neurodegeneration in the parietal lobe, fusiform gyrus, and left mid frontal gyrus,^22^ increases in WMH volume,^23,22^ and global cognitive decline.^21^ When using PET, associations of amyloid with amygdala and hippocampal volume have been observed in some studies,^24^ while associations with temporal and fusiform neurodegeneration has been observed in others.^25^ PET amyloid burden is also associated with total WMH volume^23^, and cognition,^26^ while plasma amyloid has shown associations with total brain, hippocampal,^27^ and WMH volume,^28^ as well as cognition.^29^

However, only a few studies have examined these methods together,^26,30,31^ finding discrepant associations across the different amyloid measurement modalities. For instance, PET amyloid was associated with hippocampal volume, whereas CSF amyloid showed no such association.^26^ Another study observed that while PET amyloid was associated with cognitive decline, plasma amyloid was not.^30^ Furthermore, studies have found that CSF amyloid was associated with WMHs, while PET amyloid was not associated with WMHs to the same degree.^31^

Although amyloid is associated with neurodegeneration, cerebrovascular pathology, and cognitive decline, it remains unknown if these associations remain the same using PET, CSF, or plasma to measure amyloid. Additionally, the differences observed between these three methods to measure amyloid may be associated with risk factor prevalence, particularly when using plasma.^32–34^ The current study will explore how these three amyloid measurement modalities (i.e., PET, CSF, plasma) are associated with each other and with different dementia-related outcomes including neurodegeneration, cerebrovascular pathology, and cognition.

## Methods

### Alzheimer’s Disease Neuroimaging Initiative

Data used in the preparation of this article were obtained from the Alzheimer’s Disease Neuroimaging Initiative (ADNI) database (adni.loni.usc.edu). The ADNI was launched in 2003 as a public-private partnership, led by Principal Investigator Michael W. Weiner, MD. The primary goal of ADNI has been to test whether serial MRI, positron emission tomography (PET), other biological markers, and clinical and neuropsychological assessment can be combined to measure the progression of mild cognitive impairment and early AD. The study received ethical approval from the review boards of all participating institutions. Written informed consent was obtained from participants or their study partner. Participants were selected only from all ADNI cohorts (ADNI-1, ADNI-GO, ADNI-2 and ADNI-3).

### Participants

Full participant inclusion and exclusion criteria are available at https://www.adni-info.org. All participants were between the ages of 55 and 90 at baseline. Participants were included in the present analyses if they had MRI scans, PET, CSF, and plasma amyloid measurements available. From all ADNI cohorts, 248 participants (*n* = 82 cognitive normal, CN; *n* = 140 mild cognitive impairment, MCI; *n* = 26 dementia) met the inclusion criteria. CN participants had no evidence of memory decline on the Logical Memory II subscale from the Wechsler Memory Scale and no evidence of cognitive decline on either the Mini Mental Status Examination (MMSE) or Clinical Dementia Rating (CDR) (with a score of 0 required for memory box). MCI participants had to score between 24 and 30 on the MMSE, 0.5 on the CDR with a memory box score of at least 0.5 and had abnormal scores on the Logical Memory II test. Dementia participants had to show abnormal memory function on the Logical Memory II test, an MMSE score between 20 and 26, a CDR-SB 0.5 or 1.0 and probable AD according to the National Institute of Neurological and Communicative Disorders and Stroke and the Alzheimer’s Disease and Related Disorders Association criteria.^44^

### Risk Factors

Risk factors downloaded from the ADNI website included Hachinski score, history of hypertension, BMI, alcohol usage, and tobacco smoking. Hachinski score is a score from 0-18, with scores of 4 or below suggesting a higher likelihood of AD compared to other types of dementia (e.g., vascular dementia). Since having a Hachinski score above 4 was an exclusion criterion in the ADNI^45^, included participants have Hachinski scores ranging between 0-4. History of hypertension indicates 1 = history of hypertension at any point in their life, 0 = no history of hypertension. BMI is a continuous measure calculated based on a person’s weight and height, measured as kg/m^2^. Alcohol usage indicates if the participant had a history of alcohol or substance abuse or dependence within the past 2 years (DSM-IV criteria), in which 1 = yes, 0 = no. Tobacco smoking is a yes or no question, in which 1 = yes, and 0 = no.

### Gray Matter Measurements

Hippocampal, entorhinal cortex, fusiform gyrus, temporal, ventricular, and whole brain volumes were extracted from the ADNIMERGE file provided by the ADNI. Volumetric segmentation was performed with the FreeSurfer image analysis suite available at http://surfer.nmr.mgh.harvard.edu/.

### WMH measurements

Measurements of WMHs were obtained through a previously validated WMH segmentation technique and a library of manual segmentations based on 50 ADNI participants (independent of the 248 studied here).^46,47^ WMH load was defined as the volume of all voxels identified as WMHs in the standard space (in mm^3^) and are thus normalized for head size. To achieve normal distribution, WMH volumes were log-transformed.

### Cognitive Measures

Various batteries were used to measure cognition, including the Alzheimer’s Disease Assessment Scale-Cognitive Subscale (ADAS-13), the Clinical Dementia Rating-Sum of Boxes (CDR-SB), the Mini-Mental State Examination (MMSE), the Montreal Cognitive Assessment (MoCA), the Rey Auditory Verbal Learning Test and its subscales (RAVLT learning; RAVLT immediate; RAVLT forgetting), Logical Memory - Delayed Recall Total (LDTOTAL), Trail Making Test Part B (TRABSCOR), and the Functional Assessment Questionnaire (FAQ).

### CSF Biomarkers

Baseline CSF biomarkers were collected the morning after an overnight fast, and lumbar puncture was performed according to the standard ADNI procedures. CSF amyloid concentrations were measured using the assay platforms available at the time of sample collection. ADNI 1, GO, and 2 were assayed using manual immunoassay whereas ADNI3 CSF measurements in ADNI3 were obtained using the automated Roche Elecsys immunoassay platform. All measurements underwent ADNI centralized quality control prior to release. See https://adni.loni.usc.edu/wp-content/uploads/ADNI_SteeringCommittee_Meeting_2016/6%20%5B%E2%80%A6%5D%20ADNI%20Biomarker%20Core%20v2%20%5BCompatibility%20Mode%5D.pdf for more information.

### PET Biomarkers

Amyloid PET imaging was performed using [^18F]Florbetapir (AV-45) in accordance with standardized ADNI acquisition protocols (https://adni.loni.usc.edu/wp-content/uploads/2010/05/ADNI2_PET_Tech_Manual_0142011.pdf). Cortical amyloid burden was quantified as the average standardized uptake value ratios (SUVr) across the frontal, anterior cingulate, precuneus, and parietal cortex normalized to the cerebellum. All PET imaging data underwent centralized ADNI quality control and preprocessing prior to release (https://adni.bitbucket.io/reference/docs/UCBERKELEYAV45/UCBERKELEY_AV45_Methods_11.15.2021.pdf). SUVR values were log-transformed for all statistical analyses and graphical visualization.

### Plasma Biomarkers

Baseline plasma amyloid values were obtained from the ADNI merge file. Blood plasma samples were collected in the morning after an overnight fast and prior to breakfast in accordance with the standard ADNI procedures. Plasma amyloid samples were assayed in the Biomarker core laboratory using Innogenetics AlzBia reagents on a Luminex immunoassay platform and were quantified using the AB42/40 ratio. See http://adni.loni.ucla.edu/wp-content/%20uploads/2010/11/BC_Plasma_Proteomics_Data_Primer.pdf for more information.

### Data Analysis

Analyses were performed using MATLAB R2024b. All *p*-values are reported as raw values with significance determined by false discovery rate (FDR)^48^ correction at 0.05. Independent sample t-tests were completed on the demographic information. Correlational analyses were completed to examine how amyloid measurement methods were associated with each other between the three methods (PET, CSF, plasma).

Linear regressions were completed to examine whether method of amyloid measurement (i.e., PET, CSF, plasma) would influence the association between amyloid burden and brain structure in five regions of interest (ROIs), including the hippocampus, ventricles, fusiform gyrus, entorhinal cortex, and middle temporal gyrus. Additional linear regressions examined whether regional WMH volume (i.e., total, frontal, temporal, parietal, occipital WMH volume) was associated with amyloid using the three methods. All continuous measures were z-scored and logged values were used for amyloid. Age, sex, and intracranial volume (ICV) were added to all linear regression models as covariates. The main effect of interest was the amyloid biomarker of interest (PET, CSF, plasma):

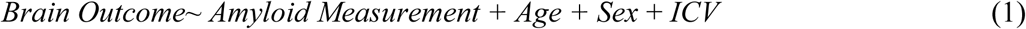

Linear regressions were also completed to examine whether each method of amyloid measurement would influence the association between cognitive scores and amyloid. Similar to the above linear regressions, all continuous measures were z-scored, logged values were used for amyloid, and age and sex were included as covariates in the model. Separate linear regressions were completed for each cognitive domain of interest. The main effect of interest was the amyloid biomarker of interest (PET, CSF, plasma).

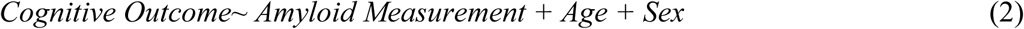

To explore whether differences in outcomes between the three measurement tools (PET, CSF, plasma) were being driven by risk factors, all analyses were repeated including the risk factors (Hachinski score, history of hypertension, BMI, alcohol usage, and tobacco smoking) as covariates in the models.

## Results

### Demographic Information

Table 2 presents the demographic and clinical characteristics of study participants. Cognitively normal participants were older than those with MCI (75.1 vs 71.6; *t*=3.4, *p*<.001), and those with MCI were younger than those with dementia (71.6 vs 75.4; *t*=-2.5, *p*=.001), CN and dementia did not differ (*p*>.05). Figure 1 presents the boxplots representing the mean and standard deviations of the three amyloid measurements. PET amyloid progressively increased from CN (1.13) to MCI (1.25) to dementia (1.46), with all groups significantly differing (*t* belongs to [-4.16 to -7.33], *p*<.001). CSF amyloid values decrease as cognitive decline progresses, and thus CN participants had greater CSF amyloid than MCI (1120.92 vs 877.75; *t*=5.22, *p*<.001) and dementia participants (1120.92 vs 689.10; *t*=5.71, *p*<.001). MCI participants also had greater CSF amyloid values than dementia participants (877.75 vs 689.10, *t*=2.78, *p*=.006). When using plasma to measure amyloid, CN participants only significantly differed from dementia participants (0.12191 vs 0.11598, *t*=2.11, *p*=.037). MCI participants did not significantly differ from CN participants (*p*=.097) or from dementia participants (*p*=.363).

**Figure 1.**
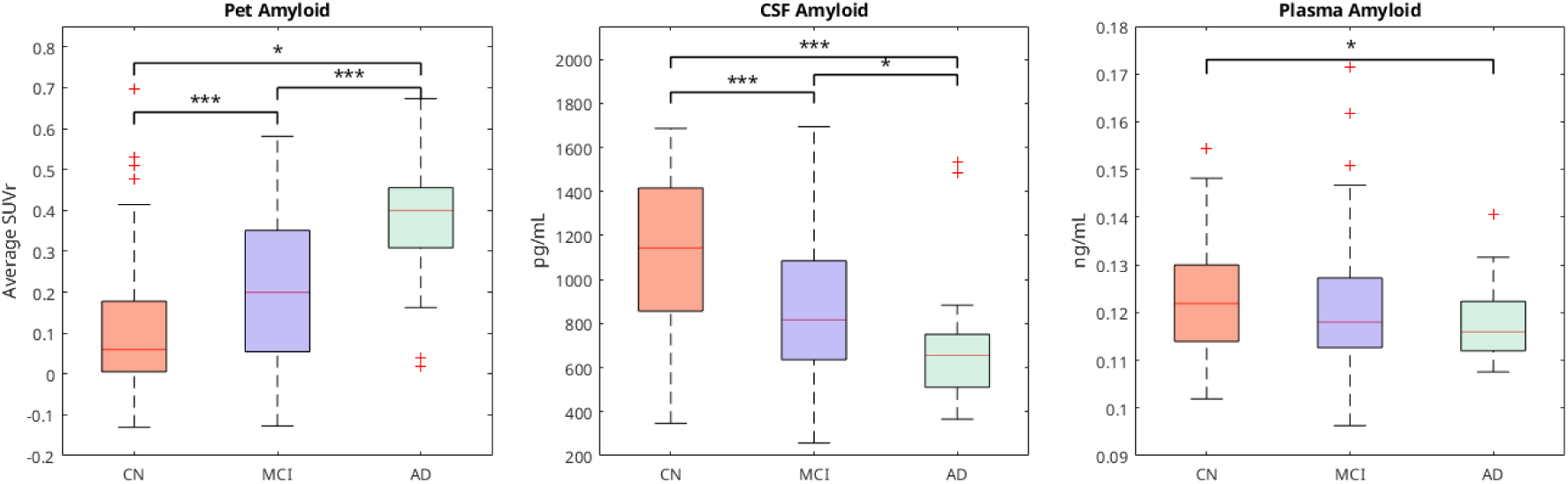
Differences in Amyloid by Biomarker Measurement. *Notes:* Group differences in PET (log transformed), CSF, and plasma biomarker levels. CN, cognitively normal; MCI, mild cognitive impairment; SUVr, standardized uptake value ratio.

**Table 2.** Descriptive statistics for participants included in this study.

|  | All Participants | CN | MCI | Dementia |
| --- | --- | --- | --- | --- |
| Participants (N) | 248 | 82 (33%) | 140 (56.5%) | 26 (10.5%) |
| Female (N) | 112 (45%) | 40 (49%) | 57 (41%) | 15 (58%) |
| Age (years) | 73.25 (7.56) | 75.14<sup>a</sup> (7.70) | 71.62<sup>c</sup> (7.21) | 75.44 (6.61) |
| Education (years) | 16.29 (2.52) | 16.41<sup>b</sup> (2.27) | 16.37 (2.61) | 15.31 (2.59) |
| PET Amyloid (SUVR) | 1.23 (0.23) | 1.13<sup>a,b</sup> (0.20) | 1.25<sup>c</sup> (0.21) | 1.46 (0.22) |
| CSF Amyloid (pg/mL) | 938.10 (356.20) | 1120.92<sup>a,b</sup> (352.69) | 877.75<sup>c</sup> (324.39) | 689.10 (275.50) |
| Plasma Amyloid (ng/mL) | 0.1207 (0.01) | 0.12191<sup>b</sup> (0.01) | 0.11797 (0.01) | 0.11598 (0.01) |

*Note. Data are number (N) and percentage or mean (standard deviation). CN = cognitively normal. MCI = mild cognitive impairment. <sup>a</sup> Significant difference ( $p < .05$ ) between CN and MCI. <sup>b</sup> Significant difference ( $p < .05$ ) between CN and Dementia. <sup>c</sup> Significant difference ( $p < .05$ ) between MCI and Dementia.*

Table 3 shows a summary of the associations between PET, CSF, and plasma amyloid, brain regions, and cognitive outcomes. PET amyloid was associated with most of the expected brain and cognition outcomes. In particular, PET amyloid was associated with all regional volumes except for the ventricles (*t* belongs to [-5.30 to -2.42], *p*<.05) as well as total (*t* = 2.15, *p*<.001) and temporal (*t* = 2.71, *p*<.001) WMHs. With respect to cognitive outcomes, PET amyloid was associated with all domains including functional activities, global cognition, memory, language, and executive functions (*t* belongs to [-8.12 to 8.20], *p*<.001). Similarly, CSF amyloid was associated with almost all expected variables. CSF amyloid was associated with all brain regions of interest (*t* belongs to [-2.93 to 4.43], *p*<.05) except for the mid temporal gyrus. Furthermore, CSF amyloid was associated with all WMH measurements (*t* belongs to [-3.76 to - 3.26], *p*<.05) except occipital WMHs. With respect to cognitive outcomes, CSF amyloid was also associated with all domains including functional activities, global cognition, memory, language, and executive functions (*t* belongs to [-6.17 to 6.69], *p*<.001). It is worth noting that while CSF amyloid was associated with more brain and cognitive regions, when significant, many of the associations with PET amyloid measures were stronger than CSF amyloid *t* scores (i.e. higher *t* values). These indicate that the associations may be stronger between PET amyloid and these specific outcomes.

**Table 3:** Linear regression outputs showing relationships between brain and cognitive measures and PET, CSF, and plasma amyloid.

|  | PET |  |  | CSF |  |  | Plasma |  |  |
| --- | --- | --- | --- | --- | --- | --- | --- | --- | --- |
|  | Estimate | tStat | pValue | Estimate | tStat | pValue | Estimate | tStat | pValue |
| **Brain Measures** |  |  |  |  |  |  |  |  |  |
| **Hippocampus** | -0.263 | -5.10 | **< .001** | 0.228 | 4.43 | **< .001** | 0.126 | 2.35 | **.020** |
| **Ventricles** | -0.008 | -0.16 | .876 | -0.144 | -2.93 | **.004** | -0.025 | -0.50 | .620 |
| **Whole Brain** | -0.104 | -3.08 | **.002** | 0.067 | 1.97 | **.049** | 0.033 | 0.95 | .343 |
| **Mid Temporal Gyrus** | -0.142 | -2.89 | **.004** | 0.081 | 1.65 | .100 | 0.036 | 0.72 | .474 |
| **Entorhinal Cortex** | -0.271 | -5.30 | **< .001** | 0.197 | 3.80 | **< .001** | 0.153 | 2.88 | **.004** |
| **Fusiform Gyrus** | -0.118 | -2.42 | **.016** | 0.130 | 2.71 | **.007** | 0.032 | 0.64 | .522 |
| **Total WMH** | 0.119 | 2.15 | **.032** | -0.202 | -3.76 | **< .001** | -0.045 | -0.81 | .419 |
| **Frontal WMH** | 0.115 | 2.07 | .040 | -0.177 | -3.26 | **.001** | -0.051 | -0.90 | .368 |
| **Parietal WMH** | 0.105 | 1.89 | .060 | -0.191 | -3.53 | **< .001** | -0.036 | -0.64 | .524 |
| **Temporal WMH** | 0.157 | 2.71 | **.007** | -0.212 | -3.75 | **< .001** | -0.120 | -2.05 | .041 |
| **Occipital WMH** | 0.022 | 0.34 | .731 | -0.120 | -1.96 | .051 | 0.033 | 0.53 | .598 |
| **Cognitive Measures** |  |  |  |  |  |  |  |  |  |
| **ADAS-13** | 0.466 | 8.20 | **< .001** | -0.363 | -6.17 | **< .001** | -0.216 | -3.44 | **< .001** |
| **CDR-SB** | 0.454 | 7.84 | **< .001** | -0.342 | -5.69 | **< .001** | -0.166 | -2.60 | **.010** |
| **MMSE** | -0.368 | -6.17 | **< .001** | 0.299 | 4.95 | **< .001** | 0.152 | 2.39 | **.018** |
| **MoCA** | -0.309 | -5.11 | **< .001** | 0.244 | 4.02 | **< .001** | 0.104 | 1.65 | .101 |
| **RAVLT forgetting** | 0.384 | 6.46 | **< .001** | -0.281 | -4.60 | **< .001** | -0.189 | -2.98 | **.003** |
| **RAVLT immediate** | -0.412 | -7.15 | **< .001** | 0.333 | 5.66 | **< .001** | 0.244 | 3.96 | **< .001** |
| **RAVLT learning** | -0.330 | -5.43 | **< .001** | 0.295 | 4.86 | **< .001** | 0.160 | 2.51 | **.013** |
| **LDTOTAL** | -0.464 | -8.12 | **< .001** | 0.392 | 6.69 | **< .001** | 0.178 | 2.80 | **.006** |
| **TRABSCOR** | 0.340 | 5.82 | **< .001** | -0.194 | -3.21 | **.001** | -0.082 | -1.32 | .188 |
| **FAQ** | 0.395 | 6.65 | **< .001** | -0.331 | -5.52 | **< .001** | -0.122 | -1.91 | .057 |

Note. WMH = White Matter Hyperintensities, PET = Positron Emission Tomography, CSF = Cerebrospinal Fluid. Bolded values are those that remained significant after FDR correction.

Of all three methods, the plasma marker for amyloid showed the fewest number of significant associations with cognition and brain outcomes, and the associations were consistently weaker compared to PET amyloid and CSF amyloid markers. Plasma amyloid was associated with only two brain regions, the hippocampus and entorhinal cortex (*t* belongs to [2.35 to 2.88], *p<.*05), and was not significantly associated with any WMH measurements. Additionally, plasma amyloid was significantly associated with the ADAS-13, CDR-SB, MMSE, RAVLT forgetting, RAVLT immediate, RAVLT total, and the LDTOAL (*t* belongs to [-3.44 to 3.96], *p*<.05) but not the MoCA, TRABSCOR, and FAQ.

### Cross-biomarker correlations

Correlational analyses were performed on the biomarker measurements, PET, CSF, and plasma, to determine if their measurements were well-correlated with each other (Figure 2). The strongest correlation was between CSF amyloid and PET amyloid (r=-0.593 *p*<.001), while CSF amyloid and plasma amyloid (r=0.338, *p*<.001) and plasma amyloid and PET amyloid were not were not correlated to the same degree (r=-0.407, *p*<.001).

**Figure 2:**
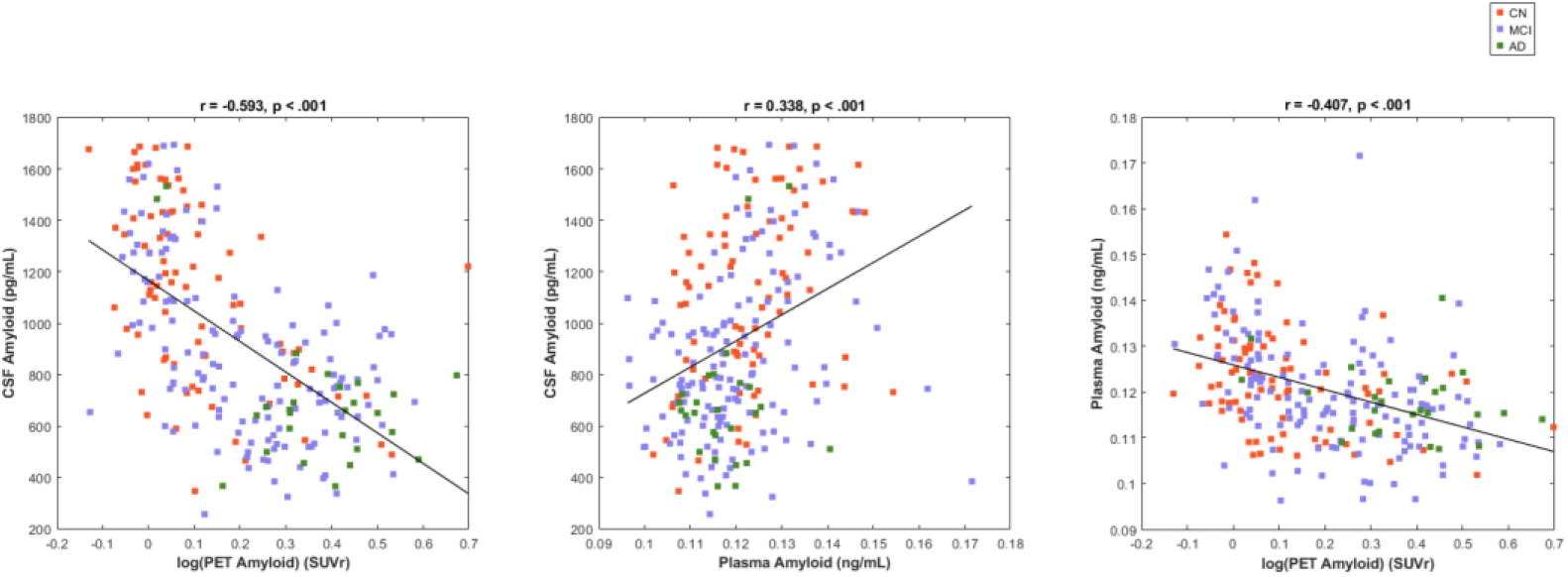
Correlation between CSF Amyloid (pg/mL), log (PET Amyloid) (SUVr), and Plasma Amyloid (ng/mL) measurements. *Notes:* A) Correlation between CSF and PET amyloid measurements. B) Correlation between CSF and Plasma amyloid measurements. C) Correlation between Plasma and PET amyloid measurements.

### Brain Regions and Biomarker Measurements

Figure 3 displays the linear regression plots examining the relationship between one region of interest (the hippocampus) and amyloid measurements for PET, CSF, and plasma. The volume of the hippocampus was significantly associated with amyloid biomarker measures using PET (*t*=-5.10, *p*<.001), CSF (*t*=4.43, *p*<.001), and plasma (*t*=2.35, *p*=.020). The regression results of the whole brain volume and the biomarker measurements were statistically significant for PET (*t*=-3.08, *p*=.002) and CSF (*t*=1.97, *p*=.049), but not plasma (*p*=.343). The entorhinal cortex was significantly associated with amyloid measurements for all methods, PET (*t*=-5.30, *p*<.001), CSF (*t*=3.80, *p*<.001), plasma (*t*=2.88, *p*=.004). The volume of the fusiform gyrus was significantly associated with amyloid biomarker measures using PET (*t*=-2.42, *p*=.016) and CSF (*t*=2.71, *p*=.007), but not plasma (*p*=.522). Ventricular volume was also only significantly associated with amyloid measurement using CSF (*t*=-2.93, *p*=.004), but not PET (*p*=.876) or plasma (*p*=.620). The volume of the middle temporal gyrus was only significantly associated with PET after FDR corrections, (*t*=-2.89, *p*=.004), but not CSF (*p*=.100) or plasma (*p*=.474).

**Figure 3:**
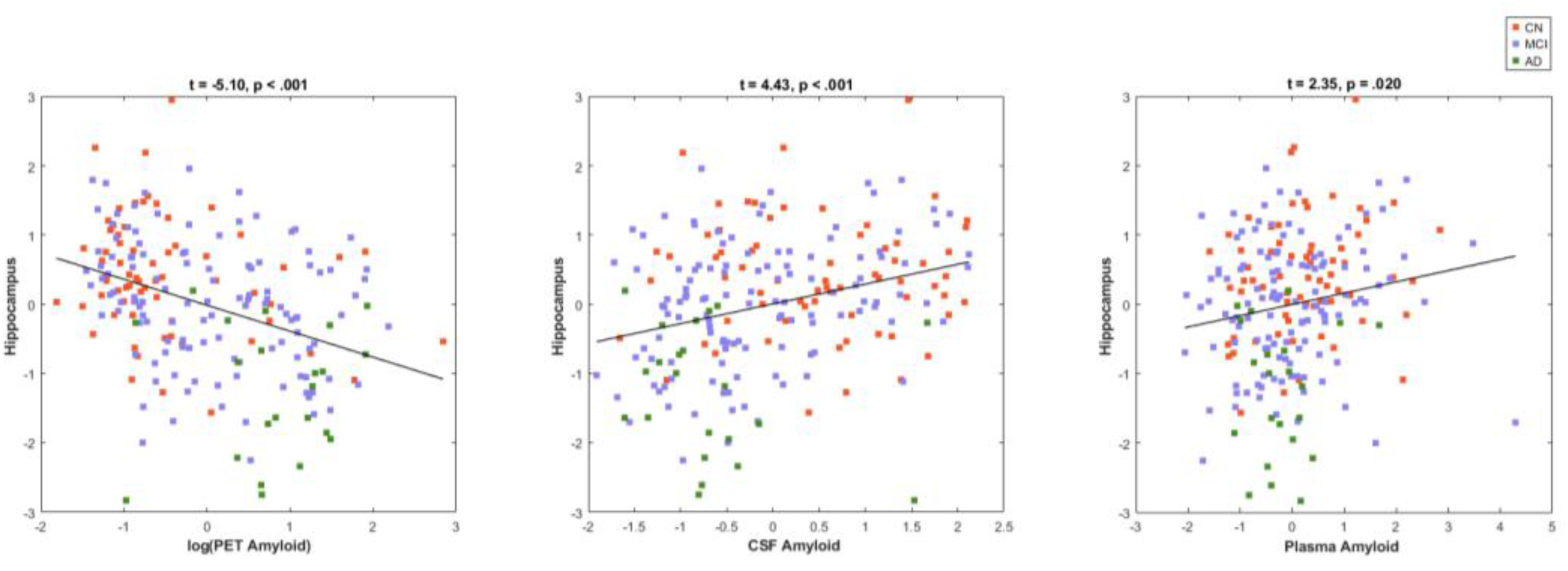
The Relationship between Hippocampal Volume and Amyloid Biomarker Measurement Methods (PET, CSF, plasma). *Notes*: PET = Positron Emission Tomography, CSF= Cerebrospinal Fluid.

### White Matter Hyperintensities and Biomarker Measurements

Figure 4 represents an example of one outcome observed for the WMH association with the three amyloid measurement techniques. Figure 4 shows a significant association between total WMH volume with CSF and PET but not plasma, and a similar trend was observed for frontal WMH. A significant association was observed between parietal WMH volume and only CSF, and a similar trend was observed for occipital WMH. A significant association was observed between temporal WMH volume and PET, CSF, and plasma.

**Figure 4:**
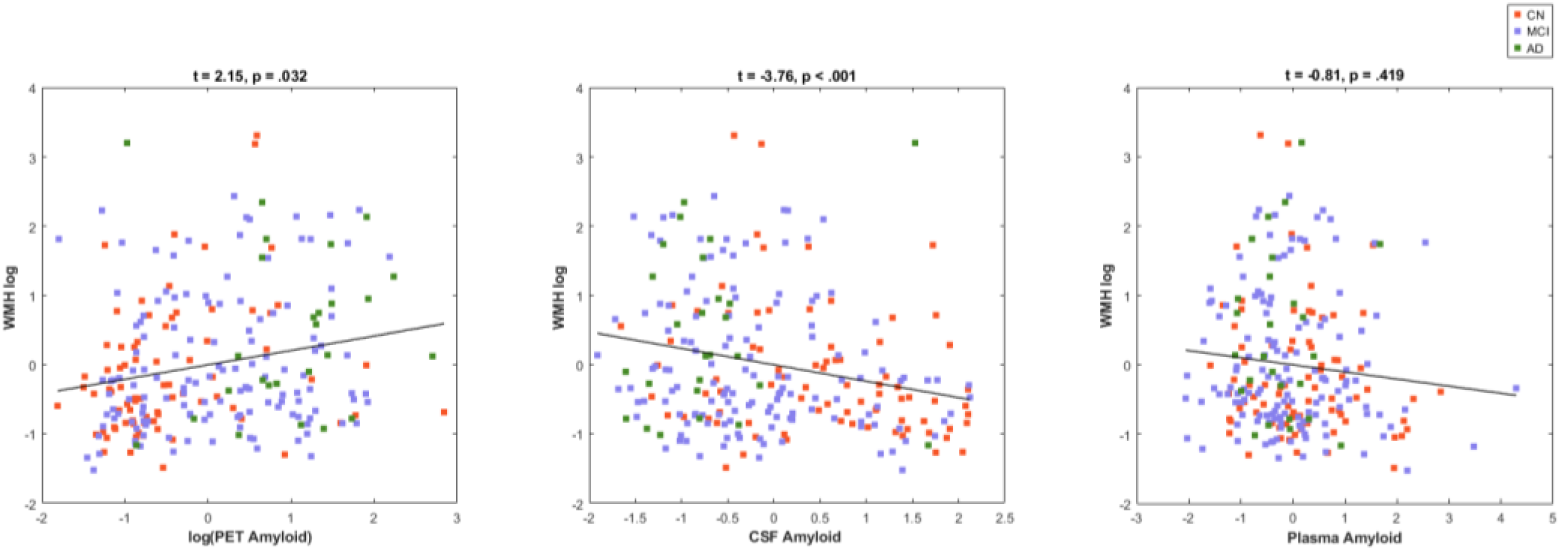
The Relationship between Total White Matter Hyperintensities and Biomarker Measurement Methods (PET, CSF, plasma) *Notes:* WMH = White Matter Hyperintensities, PET = Positron Emission Tomography, CSF = Cerebrospinal Fluid

Total WMH volume was significantly associated with amyloid biomarker measures using PET (*t*=2.15, *p*=.032) and (CSF, *t*=-3.76, *p*<.001), but was not significantly associated with amyloid measures using plasma (*p*=.419). Frontal lobe WMH volume was significantly associated with amyloid measures using CSF (*t*=-3.26, *p*=.001), but was not significantly associated with PET (*p*=.040) or plasma (*p*=.368) after FDR corrections. Parietal lobe WMH volume was significantly associated with CSF (*t*=-3.53, *p*<.001), but not with PET (*p*=.060) or plasma (*p*=.524) after FDR corrections. The occipital lobe WMH volume was not significantly associated with CSF (*p*=.051), PET (*p*=.731) or plasma (*p*=.598) after FDR corrections. Temporal lobe WMH volume was significantly associated with amyloid measures using PET (*t*=2.71, *p*=.007) and CSF (*t*=-3.75, *p*<.001), but not plasma (*p*=.041) after FDR corrections.

### Cognitive Measures and Biomarker Measurements

Figure 5 displays an example of one outcome observed between the cognitive measures and the three amyloid measurement methods. The cognitive assessment results evaluated using the MoCA revealed significant associations with PET (*t*=-5.11, *p*<.001) and CSF (*t*=4.02, *p*<.001), but not with plasma (*p*=.101). ADAS-13 showed significant relationships with all three amyloid measures, including PET (*t*=8.20, *p*<.001), CSF (*t*=-6.17, *p*<.001), and plasma (*t*=-3.44, *p*<.001). CDR-SB was significantly associated with all three biomarker measurement methods. Associations were observed between CDR-SB scores and PET (*t*=7.84, *p*<.001), CSF (*t*=-5.69, *p*<.001), and plasma, (*t*=-2.60, *p*=.010). MMSE scores were significantly associated with PET (*t*=-6.17, *p*<.001), CSF (*t*=4.95, *p*<.001), and plasma amyloid levels (*t*=2.39, *p*=.018). The RAVLT Forgetting score was significantly associated with PET (*t*=6.46, *p*<.001), CSF (*t*=-4.60, *p*<.001), and plasma (*t*=-2.98, *p*=.003). Immediate recall performance on the RAVLT was also significantly associated with PET (*t*=-7.15, *p*<.001), CSF (*t*=5.66, *p*<.001), and plasma (*t*=3.96, *p*<.001). Similarly, learning performance on the RAVLT demonstrated significant relationships with PET (*t*=-5.43, *p*<.001), CSF (*t*=4.86, *p*<.001), and plasma (*t*=2.51, *p*=.013). The delayed recall performance on the LDTOTAL was significantly associated with PET (*t*=-8.12, *p*<.001), CSF (*t*=6.69, *p*<.001) and plasma (*t*=2.80, *p*=.006). Trail Making Test Part B Score was significantly associated with PET (*t*=-5.82, *p*<.001) and CSF (*t*=-3.21, *p*=.001), however, no significant association was found with the plasma measure (*p*=.188). Performance on the FAQ was significantly associated with PET (*t*=6.65, *p*<.001) and CSF (*t*=-5.52, *p*<.001) measures, but not with plasma (*p*=.057).

**Figure 5:**
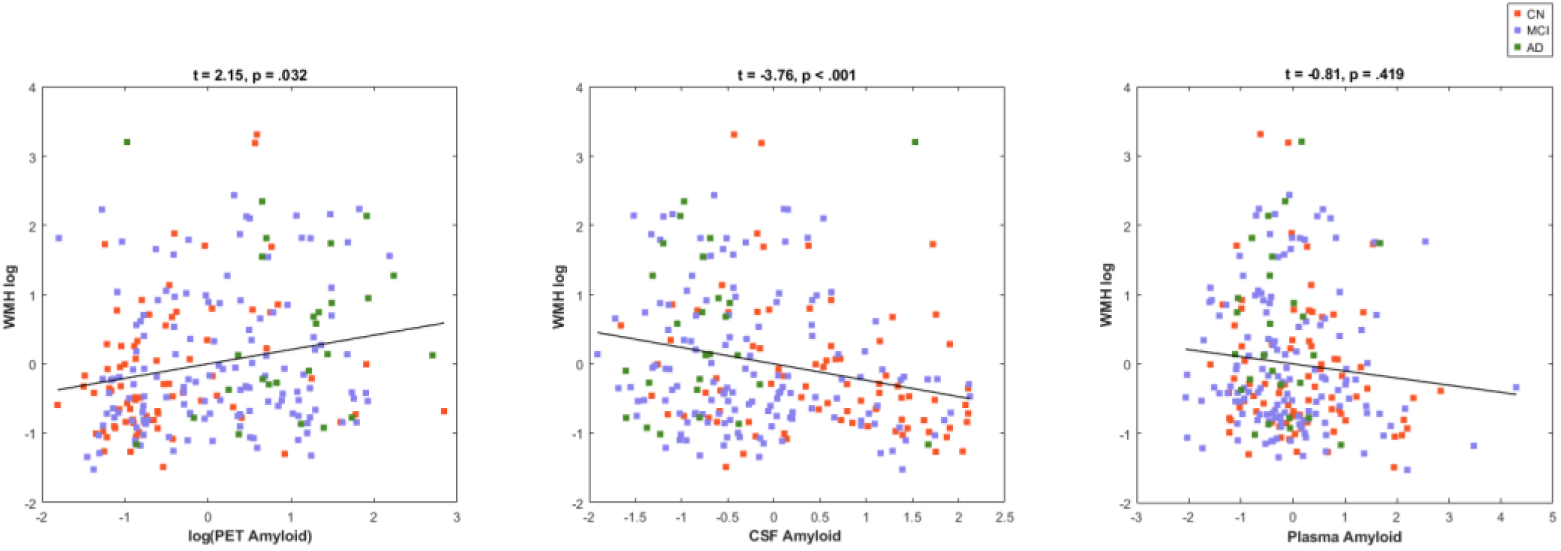
Relationship Between MoCA Scores and Amyloid Levels Measured by PET, CSF, and Plasma. *Notes:* PET = Positron Emission Tomography, CSF= Cerebrospinal Fluid, MOCA = Montreal Cognitive Assessment

Additional analyses were completed, controlling for risk factors in all models. When risk factors were added, results remained the same in terms of effect size and significance.

## Discussion

Preceding studies investigating AD biomarkers have reported conflicting findings on the relationship between amyloid biomarkers, brain structural changes and cognition.^35^ One potential reason for these conflicting results may use of different methods to measure amyloid (i.e., PET, CSF, or plasma). Although these three methods are assumed to reflect the same underlying pathological process of amyloid deposition, they capture distinct aspects of amyloid biology. PET imaging primarily quantifies fibrillar amyloid plaque burden in cortical brain tissue,^53,54^ CSF amyloid concentrations reflect soluble amyloid, which are inversely related to plaque accumulation,^54^ and plasma amyloid measures represent a combination of central and peripheral amyloid production and clearance mechanisms and thus may show weaker associations with brain amyloid burden^55^. Therefore, research is needed to explore if the contradictory findings in current research arise from comparing results that use different methods to quantify amyloid. The current study found that PET, CSF, and plasma were not strongly correlated with each other. Furthermore, the association between these amyloid biomarker measurements and MRI outcomes (regional gray matter and WMH volumes) as well as the association between these amyloid measurements and cognitive measures differed depending on the method of measurement. Lastly, we found that risk factors did not influence biomarker measurement methods in their association with MRI outcomes or cognitive measures.

Consistent with prior research, our results show that amyloid levels measured using PET and CSF significantly differ between CN, MCI, and AD.^56,57^ When using plasma to measure amyloid, however, there was no significant difference between CN and MCI participants, nor was there a difference between MCI and participants with AD. Only the comparison between CN and AD showed a significant difference in plasma-based amyloid measurements. This finding indicates the specific plasma method and assay employed here may not be sensitive enough to differentiate between cognitively normal older adults and those with prodromal dementia (i.e., MCI) and may thus have limited utility for early AD staging. Other assay methods have observed high sensitivity of plasma methods^58,59^. For example, Ashton and colleagues observed higher sensitivity than our study in their plasma measurement method using ultrasensitive single molecule array (Simoa) assays^58^. Together these findings suggest that conventional immunoassays (such as the one used in the current study), show lower diagnostic accuracy than those with ultrasensitive platforms^58,59^. Given the limited correlations and associations using the plasmas method employed here, future work should consider alternative assays to obtain higher accuracy.

In the current study, we found that CSF and PET were moderately correlated, PET and plasma were moderately correlated, and CSF and plasma were only weakly correlated with each other. These moderate-to-weak correlations between the three methods may help explain the contradictory findings reported across prior studies.^50–52^ This inconsistency has important implications for the development and clinical use of AD-related pharmaceutical drugs. For example, the two FDA-approved immunotherapy drugs lecanemab and donanemab reduce amyloid plaques.^11^ Prior to prescribing these medications, clinicians may use a PET scan or CSF analysis to confirm amyloid pathology. However, if these methods do not quantify amyloid burden in the same way, a patient’s amyloid status may vary depending on the method used. Such discrepancies may lead to misclassifying patients which could influence treatment decisions. In addition, it is important to note that some studies have found that CSF amyloid may detect cortical amyloid accumulation earlier than PET amyloid,^60,61^ a consideration that may be relevant when evaluating whether to initiate treatments or interventions with memory complaints begin or alongside early signs of AD symptomology.

The relationships between PET, CSF, and plasma-based amyloid measures and MRI outcomes also differed. As ventricular and hippocampal volume are the two most reported brain regions that experience neurodegeneration in AD,^62,63^ it is notable that their associations with amyloid varied by modality. More specifically, amyloid measured by CSF was significantly associated with ventricular volume, while PET and plasma amyloid were not. Hippocampal volume, however, was significantly associated with amyloid when measured by each of the three methods. Most associations between CSF and MRI outcomes were significant, excluding the mid-temporal gyrus, and CSF was associated with all WMH regions except occipital WMHs. While PET was associated with the same number of brain regions, it was only associated with two WMH regions, consistent with previous literature showing mixed relationships between amyloid PET and WMH burden^64^. Plasma amyloid was only associated with the volumes of the hippocampus and entorhinal cortex, but no other GM or WMH regions. These findings show that, consistent with previous studies, PET amyloid might be more strongly associated with GM atrophy, whereas CSF amyloid is more strongly associated with WMH burden, and consistently weaker associations are observed with plasma.^65,66^

Similar to the discrepant relationships observed with the MRI outcomes, we found that the associations between PET, CSF, and plasma, with cognitive measures also differed^67^. Multiple cognitive measures including the ADAS-13, CDR-SB, and MMSE were significantly correlated with all three amyloid measurements. In contrast, other measures, such as the MoCA, were only significantly associated with PET and CSF. This pattern aligns with our previous suggestion that plasma amyloid may be less sensitive to early changes in prodromal AD than PET and CSF. Among the cognitive tests examined, the MoCA is considered the most sensitive to MCI.^68,69^ Taken together, the absence of plasma-based differences between CN and MCI or between MCI and dementia, combined with the lack of association between plasma and MoCA scores, suggests that plasma may not be responsive enough to detect early-stage cognitive changes.^68^

Amyloid PET is considered the gold standard for measuring amyloid deposition, while CSF biomarkers also provide a highly validated biomarker for in vivo pathological classification^67^. Nevertheless, both approaches have several limitations. For example, PET scans are expensive and not widely accessible, which may limit applicability for individuals who need them.^70^ Similarly, CSF collection requires an invasive lumbar puncture, a procedure that many older adults may be unwilling to complete.^71^ Plasma based amyloid measurements, however, are less expensive than PET and considerably less invasive than CSF, which has increased interest in their use among AD-related researchers and clinicians.^72^ However, more studies are needed to ensure that plasma is a valid and reliable method to measure amyloid in individuals at earlier disease stages. Some studies have reported concordance and high diagnostic accuracy between plasma biomarkers with traditional biomarkers (e.g., PET, CSF),^73,74^ however, our results did not align with these findings. A potential reason for these differing results may include the methodological approach to quantify amyloid. For example, other studies used an amyloid threshold,^73,74^ while our study analyzed amyloid as a continuous measure. For example, using amyloid-positive thresholds for PET, CSF, and plasma to determine if a participant meets the criteria for AD rather than using amyloid values as a continuous variable, may lead to greater correlations between the measurement methods. These studies, however, did not report the specific plasma amyloid threshold used, and utilized amyloid ratios.^73^ Furthermore, Altomare and colleagues state that the threshold used in their study might not generalize to other studies due to differences between samples.^73^

There are a few limitations of the current study that should be noted. Since limited longitudinal data was available for the subset of participants that had all three amyloid measures of interest, our study analyzed baseline data from the ADNI database, therefore, we cannot assess longitudinal effects, nor establish causation. Furthermore, findings from the ADNI database might not generalize to other populations, as most ADNI participants are white and well-educated, and not representative of the general population^75^.

As researchers aim to diagnose early AD using biomarkers such as amyloid and tau, the choice of measurement modality becomes increasingly important. In this study, PET, CSF, and plasma, demonstrated only modest correlations, raising the concern of possible inconsistent amyloid classification across these methods. Although PET and CSF were more strongly correlated with each other than either was with plasma, plasma assays offer greater feasibility and accessibility than PET and CSF. Our findings also suggest that these three different modalities have varying levels of associations with structural brain changes (both neurodegeneration and WMHs) and cognitive measures. These differing associations were not influenced by risk factors. Currently, these results suggest that PET and CSF measures correlated with brain and cognitive changes better than plasma, and that the plasma method used in this study is less responsive to early changes in cognitive decline.

## Data Availability

All data produced are available online at adni.loni.usc.edu

## Acknowledgments

Data collection and sharing for this project was funded by the Alzheimer’s Disease Neuroimaging Initiative (ADNI) (National Institutes of Health Grant U01 AG024904) and DOD ADNI (Department of Defense award number W81XWH-12-2-0012). ADNI is funded by the National Institute on Aging, the National Institute of Biomedical Imaging and Bioengineering, and through generous contributions from the following: AbbVie, Alzheimer’s Association; Alzheimer’s Drug Discovery Foundation; Araclon Biotech; BioClinica, Inc.; Biogen; Bristol-Myers Squibb Company; CereSpir, Inc.; Cogstate; Eisai Inc.; Elan Pharmaceuticals, Inc.; Eli Lilly and Company; EuroImmun; F. Hoffmann-La Roche Ltd and its affiliated company Genentech, Inc.; Fujirebio; GE Healthcare; IXICO Ltd.; Janssen Alzheimer Immunotherapy Research & Development, LLC.; Johnson & Johnson Pharmaceutical Research & Development LLC.; Lumosity; Lundbeck; Merck & Co., Inc.; Meso Scale Diagnostics, LLC.; NeuroRx Research; Neurotrack Technologies; Novartis Pharmaceuticals Corporation; Pfizer Inc.; Piramal Imaging; Servier; Takeda Pharmaceutical Company; and Transition Therapeutics. The Canadian Institutes of Health Research is providing funds to support ADNI clinical sites in Canada. Private sector contributions are facilitated by the Foundation for the National Institutes of Health (https://www.fnih.org). The grantee organization is the Northern California Institute for Research and Education, and the study is coordinated by the Alzheimer’s Therapeutic Research Institute at the University of Southern California. ADNI data are disseminated by the Laboratory for Neuro Imaging at the University of Southern California.

## Consent Statement

Written informed consent was obtained from participants or their study partner.

